# Booster Vaccination with SARS-CoV-2 mRNA Vaccines and Myocarditis Risk in Adolescents and Young Adults: A Nordic Cohort Study of 8.9 Million Residents

**DOI:** 10.1101/2022.12.16.22283603

**Authors:** Anders Hviid, Tuomo A. Nieminen, Nicklas Pihlström, Nina Gunnes, Jesper Dahl, Øystein Karlstad, Hanne Løvdal Gulseth, Anders Sundström, Anders Husby, Jørgen Vinsløv Hansen, Rickard Ljung, Petteri Hovi

## Abstract

**Importance:** The SARS-CoV-2 mRNA vaccines are associated with an increased risk of myocarditis. This association appears to be strongest in male adolescents and younger males and after the second dose. Few studies have evaluated the association after booster doses.

**Objective:** To evaluate the risk of myocarditis following SARS-CoV-2 mRNA booster vaccination in 12-to-39-year-olds.

**Design:** A multinational cohort study using nationwide register data.

**Setting:** Denmark, Finland, Norway, and Sweden.

**Participants:** Cohorts comprising all 8.9 million individuals residing in each of the four countries, born 1982-2009, and alive at start of study on December 27, 2020, without a previous diagnosis of myocarditis or pericarditis or laboratory-confirmed SARS-CoV-2 infection.

**Exposures:** The 28-day acute risk periods following the second and third dose of BNT162b2 and mRNA-1273, respectively, in a homologous schedule defines the exposures of interest.

**Main Outcomes and Measures:** Cohort participants were followed until an inpatient diagnosis of myocarditis, loss to follow-up, or end of study (latest data availability in each country), whichever occurred first. In each of the four countries, Poisson regression was used to estimate adjusted incidence rate ratios (IRRs) of myocarditis, with associated 95% confidence intervals (CIs), according to vaccination status. Country-specific results were combined in meta-analyses.

**Results:** A total of 8.9 million residents were followed for 12,271,861 person-years. We identified 1533 cases of myocarditis. In 12-to-39-year-old males, the 28-day acute risk period following the third dose of BNT162b2 or mRNA-1273 was associated with an increased incidence rate of myocarditis compared to the post-acute risk period 28 days or more after a second homologous dose (IRR, 2.08 [95% CI, 1.31 to 3.33] and 8.89 [95% CI, 2.26 to 35.03], respectively). The corresponding incidence rates following the third dose of BNT162b2 and mRNA-1273 were 0.86 and 1.95, respectively, within 28 days of follow-up among 100,000 individuals.

**Conclusions and Relevance:** Our results suggest that a booster dose is associated with increased myocarditis risk in male adolescents and young male adults.

**KEY POINTS:** *Question:* Is a first booster dose of SARS-CoV-2 messenger RNA (mRNA) vaccine associated with increased risk of myocarditis in male adolescents and young male adults?

*Findings:* In a cohort study of 8.9 million residents in Denmark, Finland, Norway, and Sweden, a booster dose of BNT162b2 or mRNA-1273 vaccine was associated with increased risk of myocarditis in 12-to-39-year-old males, with incidence rates 0.86 and 1.95, respectively, within 28 days of follow-up per 100,000 vaccinated individuals.

*Meaning:* A booster dose with a SARS-CoV-2 mRNA vaccine is associated with increased myocarditis risk in male adolescents and young male adults. Compared to after a primary course, the risk appears attenuated.

---

Myocarditis is a rare adverse event following vaccination with the two available SARS-CoV-2 mRNA vaccines BNT162b2 (BNT) and mRNA-1273 (MOD). Previous studies have shown that the association is strongest in individuals below the age of 40 years, in males, and after the second dose.^1–4^ Currently available evidence also suggests increased myocarditis risk associated with booster doses.^5–8^ In a self-controlled case series study from the UK, relative risks of 1.72 and 2.64 were observed for the 28-day risk period following a BNT or MOD booster dose vs. reference periods, respectively.^6^ These relative risk estimates were lower than those observed for the 28-day period following second doses of BNT and MOD. However, the absolute risk of myocarditis was very low in this setting, with 1–2 excess cases per 1,000,000 booster vaccinations among individuals 13 years or older.^6^ Increased risks after the booster dose have also been observed in a French case-control study, in a US cohort of Kaiser Permanente Southern California members and in a Canadian cohort.^5,7,8^ In all three studies, the relative risks associated with the booster dose were lower than the relative risks associated with the second dose.

In the current study based on nationwide cohorts comprising the adolescent and young adult populations in the four largest Nordic countries, we build on our previous comprehensive evaluation of one- and two-dose mRNA schedules and myocarditis risk,^4^ and add significantly to the evidence on the associations between mRNA vaccine booster doses and risk of myocarditis.

## METHODS

### Setting and Data Sources

We constructed country-specific cohorts taking advantage of the near real-time nationwide register data available in the Nordic countries (Denmark, Finland, Norway, and Sweden). Our study cohorts comprised all individuals residing in the four countries, born from 1982 to 2009, and alive at study start on December 27, 2020. Using unique national person identifiers, we linked information on positive polymerase chain reaction (PCR) SARS-CoV-2 test results, SARS-CoV-2 vaccinations, myocarditis diagnoses in the hospital setting, and potential confounders from nationwide registers.^4^ Individuals with at least one primary or secondary diagnosis of either myocarditis or pericarditis (International Statistical Classification of Diseases and Related Health Problems, 10^th^ Revision [ICD-10] codes: I40.0, I40.1, I40.8, I40.9, I41.1, I41.8, I51.4, I30.0, I30.1, I30.8, I30.9, I32.8) in either inpatient or outpatient hospital care in the period from January 1, 2017, until the study start, were excluded from the study. Individuals with a laboratory-confirmed SARS-CoV-2 infection before the study start were also excluded from the study.

### Vaccinations and infections

Individual-level information on dates and types of all SARS-CoV-2 vaccinations (both mRNA vaccines, viral vector vaccines, and the protein-based vaccine) was obtained from national vaccination registers.^4^ The Nordic countries have used mRNA vaccines extensively. Primary courses (two doses) were largely administered to the general adult population from June 2021 to September 2021, while the first booster doses were administered from November 2021 to February 2022. In Sweden, the use of MOD was limited to residents 30 years of age or older from October 6, 2021. The next day, the Finnish authorities recommended that only BNT would be used for males below 30 years. In Norway, all children below 18 years were recommended BNT, and from October 6, 2021, all men below 30 years were advised to prefer BNT, and from January 12, 2022, the recommendation was extended to women below 30 years. Denmark did not provide specific recommendations.

Individual-level information on dates of positive PCR SARS-CoV-2 tests from national surveillance databases was obtained.^4^ In Denmark, until February 2022, testing was easily accessible, free of charge, and available to all, regardless of indication. The situation was similar in Norway until January 2022, when only unvaccinated, partially vaccinated, and symptomatic adults were recommended testing. After initial availability issues during spring 2020, the situation was similar in Finland until August 26, 2021, when testing decreased. In Sweden, the testing capacities were more limited and reserved primarily for symptomatic cases. In February 2022, the testing capacity was downscaled throughout the Nordic countries.

### Myocarditis

Individual-level information on dates of hospital admission for myocarditis was obtained from national hospital registers.^4^ A myocarditis outcome was defined as the first occurrence of a main or secondary myocarditis diagnosis (ICD-10 codes: I40.0, I40.1, I40.8, I40.9, I41.1, I41.8, or I51.4) at discharge from inpatient hospital care. The outcome date was the date of hospital admission.

### Statistical analysis

The country-specific cohorts were followed from study start on December 27, 2020, until a myocarditis diagnosis, vaccination with a fourth dose, vaccination with any other SARS-CoV-2 vaccines than the mRNA vaccines or the AstraZeneca vaccine AZD1222 (AZD), a laboratory-confirmed SARS-CoV-2 infection, or loss to follow-up due to death, emigration, or study end (latest country-specific date of data availability: Denmark, August 31, 2022; Finland, September 1, 2022; Norway, July 31, 2022; Sweden, June 30, 2022), whichever occurred first. If an individual had a myocarditis outcome on the same day as vaccination, the vaccination was assumed to have occurred first. Individuals contributed with unvaccinated follow-up until their first SARS-CoV-2 vaccination. Following each dose, individuals contributed time in two non-overlapping periods, a 28-day acute risk period of interest starting on day 0 (day of vaccination) and ending on day 27, followed by a post-acute risk period starting on day 28 and ending when another dose was received or follow-up ended. If and when a new dose was received, a new 28-day acute risk period was entered and followed by the post-acute risk period. Vaccination status was classified by the type- and dose-specific schedule received. The most prevalent booster schedules were the homologous schedules, BNT1BNT2BNT3 (three doses of BNT) and MOD1MOD2MOD3 (three doses of MOD). The resulting counts of myocarditis and person-years of follow-up were aggregated according to vaccination status and potential confounders.

In each country, we used Poisson regression to estimate adjusted incidence rate ratios (IRRs) of myocarditis, with associated 95% confidence intervals (CIs). We estimated IRRs comparing a) the 28-day acute risk period after the booster dose to unvaccinated follow-up, b) the 28-day acute risk period after the booster dose to the post-acute risk period 28 days or more after the second dose, and c) the 28-day acute risk period after the second dose to unvaccinated follow-up. We adjusted for sex, attained age in 2021 (12– 15, 16–24, 25–29, or 30–39 years), calendar period (December 2020 through February 2021, March and April 2021, May and June 2021, July and August 2021, September and October 2021, November and

December 2021, January and February 2022, March and April 2022, May and June 2022, or July 2022 onwards), and vaccination priority group (country-specific definition, e.g., frontline personnel and high-risk individuals). Each regression model also included terms corresponding to the interaction between vaccination status and sex, allowing for sex-specific effects of vaccination status. Separate regression analyses were performed for the following age strata: 12–15, 16–24, 25–39, and 12–39 years.

Country-specific results were combined in a meta-analysis using random-effects models implemented in the *mixmeta* package in R^9,10^, which provided pooled log IRR estimates and standard errors. An IRR estimate from a specific country was included in the meta-analysis if its standard error was positive and finite, which excluded IRR estimates resulting from zero events in either the risk or comparison period. We used a normal approximation to derive 95% CIs for the pooled log IRR estimates. We also calculated pooled incidence rates of myocarditis by age, sex, and vaccination status by using the sum of events and person-years from all the countries and derived 95% CIs by assuming a Poisson distribution on the event counts. Incidence rates are presented as the number of myocarditis cases during 28 days among 100,000 individuals.

## RESULTS

### Description of the Nordic cohorts

After the exclusion of 285,927 individuals before study start (278,671 due to COVID-19 and 7,256 due to previous myocarditis or pericarditis), we included a total of 8,859,339 12-to-39-year-olds (4,291,312 females, 48%) in four country-specific cohorts (1,944,196 from Denmark, 1,891,033 from Finland, 1,833,908 from Norway, and 3,190,202 from Sweden). From study start on December 27, 2020, and until country-specific study end, 3,119,286 had only received a primary course of homologous mRNA vaccines (2,650,414 [85%] in the BNT1BNT2 schedule and 468,872 [15%] in the MOD1MOD2 schedule), and 2,384,885 had received a homologous mRNA vaccine booster dose (2,062,026 [86%] in the BNT1BNT2BNT3 schedule and 322,859 [14%] in the MOD1MOD2MOD3 schedule). A total of 1,595,443 (18%) were unvaccinated, and 1,759,725 (20%) were only partially vaccinated or had received heterologous schedules. Uptake of both primary courses and booster doses was similar across countries and was generally high even in these younger age groups (Table 1). BNT was the most widely used vaccine of the two mRNA vaccines.

**Table 1:** Vaccination status at study end among 12-to-39-year-olds in Denmark, Finland, Norway, and Sweden.

|  |  | Vaccination status at study end (% among all 12-39-year-olds)<sup>1</sup> |  |  |  |  |  |
| --- | --- | --- | --- | --- | --- | --- | --- |
| Sex | All 12-to-39-year-olds | Unvaccinated | BNT1BNT2 | BNT1BNT2BNT3 | MOD1MOD2 | MOD1MOD2MOD3 | All other schedules<sup>2</sup> |
| Denmark |  |  |  |  |  |  |  |
| All | 1944196 (100.0%) | 294465 (15.1%) | 563534 (29.0%) | 643656 (33.1%) | 107793 (5.5%) | 215176 (11.1%) | 119572 (6.2%) |
| Females | 951246 (48.9%) | 133764 (6.9%) | 269126 (13.8%) | 331012 (17.0%) | 51612 (2.7%) | 104755 (5.4%) | 60977 (3.1%) |
| Males | 992950 (51.1%) | 160701 (8.3%) | 294408 (15.1%) | 312644 (16.1%) | 56181 (2.9%) | 110421 (5.7%) | 58595 (3.0%) |
| Finland |  |  |  |  |  |  |  |
| All | 1891033 (100.0%) | 344860 (18.2%) | 626856 (33.1%) | 416368 (22.0%) | 112551 (6.0%) | 29242 (1.5%) | 361156 (19.1%) |
| Females | 913413 (48.3%) | 141107 (7.5%) | 283733 (15.0%) | 216839 (11.5%) | 69806 (3.7%) | 20332 (1.1%) | 181596 (9.6%) |
| Males | 977620 (51.7%) | 203753 (10.8%) | 343123 (18.1%) | 199529 (10.6%) | 42745 (2.3%) | 8910 (0.5%) | 179560 (9.5%) |
| Norway |  |  |  |  |  |  |  |
| All | 1833908 (100.0%) | 246360 (13.4%) | 367609 (20.0%) | 322005 (17.6%) | 76072 (4.1%) | 32836 (1.8%) | 789026 (43.0%) |
| Females | 889347 (48.5%) | 105918 (5.8%) | 176039 (9.6%) | 171236 (9.3%) | 35695 (1.9%) | 16625 (0.9%) | 383834 (20.9%) |
| Males | 944561 (51.5%) | 140442 (7.7%) | 191570 (10.4%) | 150769 (8.2%) | 40377 (2.2%) | 16211 (0.9%) | 405192 (22.1%) |
| Sweden |  |  |  |  |  |  |  |
| All | 3190202 (100.0%) | 709758 (22.2%) | 1092415 (34.2%) | 679997 (21.3%) | 172456 (5.4%) | 45605 (1.4%) | 489971 (15.4%) |
| Females | 1537306 (48.2%) | 307478 (9.6%) | 503763 (15.8%) | 368354 (11.5%) | 75322 (2.4%) | 22928 (0.7%) | 259461 (8.1%) |
| Males | 1652896 (51.8%) | 402280 (12.6%) | 588652 (18.5%) | 311643 (9.8%) | 97134 (3.0%) | 22677 (0.7%) | 230510 (7.2%) |

<sup>1</sup>Status at country-specific study end (Denmark: August 31, 2022; Finland: September 1, 2022; Norway: July 31, 2022; Sweden: June 30, 2022)
<sup>2</sup>Comprises BNT1, MOD1, AZD1 and two- and three-dose heterologous schedules.

### Comparisons according to booster vaccination status and sex

During 12,271,861 person-years of follow-up, we identified 1,533 cases (270 females, 18%) of myocarditis, corresponding to an incidence rate of 12.5 cases during 100,000 person-years of follow-up. Among males, we observed increased incidence rates comparing the 28-day acute risk period after a booster schedule of BNT1BNT2BNT3 or MOD1MOD2MOD3 to unvaccinated follow-up (Table 2). IRRs for the 28-day risk period were attenuated for the booster dose compared to the second dose, although confidence intervals overlapped. Among males, the IRR for MOD1MOD2MOD3 was 6.47 (95% CI, 2.09 to 20.01), whereas the IRR for MOD1MOD2 was 12.56 (95% CI, 9.42 to 16.73) (Table 2). Furthermore, the IRR for BNT1BNT2BNT3 was 2.21 (95% CI, 1.37 to 3.57), whereas the IRR for BNT1BNT2 was 2.86 (95% CI, 2.23 to 3.68) (Table 2).

**Table 2:** Association between mRNA vaccination schedules and myocarditis among 12-to-39-year-olds in Denmark, Finland, Norway, and Sweden.

| Age | Sex<sup>1</sup> | Events | Person-years of follow-up | Incidence rate – cases in 28 days among 100,000 individuals | Incidence rate ratio (95% CI)<sup>2</sup> | Contributing countries<sup>3</sup> |
| --- | --- | --- | --- | --- | --- | --- |
| BNT1BNT2 acute 28-day risk period vs unvaccinated follow-up |  |  |  |  |  |  |
| 12-39 | F | 20 | 521160.09 | 0.28 | 2.56 (1.54 to 4.24) | DK,FI,SE |
| 12-15 | F | 2 | 45827.46 | 0.28 | 12.56 (1.67 to 94.22) | FI,SE |
| 16-24 | F | 8 | 160806.33 | 0.36 | 3.12 (1.36 to 7.17) | DK,FI,SE |
| 25-39 | F | 10 | 307206.31 | 0.24 | 2.20 (1.09 to 4.46) | DK,FI,SE |
| 12-39 | M | 96 | 551252.53 | 1.34 | 2.86 (2.23 to 3.68) | DK,FI,NO,SE |
| 12-15 | M | 9 | 55890.23 | 1.22 | 3.59 (1.52 to 8.51) | DK,FI,SE |
| 16-24 | M | 60 | 170560.15 | 2.70 | 4.13 (2.90 to 5.88) | DK,FI,NO,SE |
| 25-39 | M | 27 | 323930.60 | 0.64 | 1.77 (1.14 to 2.76) | DK,FI,NO,SE |
| BNT1BNT2BNT3 acute 28-day risk period vs unvaccinated follow-up |  |  |  |  |  |  |
| 12-39 | F | 4 | 167827.82 | 0.15 | 3.14 (0.79 to 12.57) | DK,FI |
| 12-39 | M | 21 | 186849.06 | 0.86 | 2.21 (1.37 to 3.57) | DK,FI,NO,SE |
| BNT1BNT2BNT3 acute 28-day risk period vs BNT1BNT2 post-acute risk period |  |  |  |  |  |  |
| 12-39 | F | 4 | 167827.82 | 0.15 | 3.99 (0.41 to 38.64) | DK,FI |
| 12-39 | M | 21 | 186849.06 | 0.86 | 2.08 (1.31 to 3.33) | DK,FI,NO,SE |
| 16-24 | M | 11 | 48997.33 | 1.59 | 2.67 (1.38 to 5.17) | DK,FI,SE |
| 25-39 | M | 10 | 132327.03 | 0.58 | 1.92 (0.97 to 3.81) | DK,FI,NO,SE |
| MOD1MOD2 acute 28-day risk period vs unvaccinated follow-up |  |  |  |  |  |  |
| 12-39 | F | 5 | 102917.95 | 0.37 | 3.82 (1.52 to 9.60) | DK,FI,NO,SE |
| 25-39 | F | 4 | 55866.20 | 0.55 | 4.22 (1.47 to 12.12) | DK,FI,NO,SE |
| 12-39 | M | 71 | 79227.55 | 6.87 | 12.56 (9.42 to 16.73) | DK,FI,NO,SE |
| 16-24 | M | 29 | 15655.35 | 14.20 | 15.04 (9.57 to 23.63) | DK,FI,NO,SE |
| 25-39 | M | 38 | 54509.16 | 5.34 | 10.21 (6.78 to 15.38) | DK,FI,NO,SE |
| MOD1MOD2MOD3 acute 28-day risk period vs unvaccinated follow-up |  |  |  |  |  |  |
| 12-39 | M | 4 | 8214.30 | 1.95 | 6.47 (2.09 to 20.01) | DK,SE |
| MOD1MOD2MOD3 acute 28-day risk period vs MOD1MOD2 post-acute risk period |  |  |  |  |  |  |
| 12-39 | M | 4 | 8214.30 | 1.95 | 8.89 (2.26 to 35.03) | DK,SE |

| Age | Sex<sup>1</sup> | Events | Person-years of follow-up | Incidence rate – cases in 28 days among 100,000 individuals | Incidence rate ratio (95% CI)<sup>2</sup> | Contributing countries<sup>3</sup> |
| --- | --- | --- | --- | --- | --- | --- |
| 25-39 | M | 3 | 7721.19 | 1.52 | 8.06 (1.81 to 35.94) | DK,SE |

<sup>1</sup>Biological sex. F, female; M, male.
<sup>2</sup>Adjusted for age (attained age in 2021: 12–15, 16–24, 25–29, or 30–39 years of age), calendar period (December 2020 through February 2021, March and April 2021, May and June 2021, July and August 2021, September and October 2021, November and December 2021, January and February 2022, March and April 2022, May and June 2022, or July 2022 onwards), and vaccination priority group (country-specific definition, e.g., frontline personnel and high-risk individuals).
<sup>3</sup>Countries contributing information to the meta-analyses of the incidence rate ratios. The reason for not participating in an analysis was no events in either the exposed group or the reference group. Incidence rates were calculated using follow-up from all countries. DK, Denmark; FI, Finland; NO, Norway; SE, Sweden.

We also compared incidence rates after booster doses to the incidence rates preceding it. In the comparison between the 28-day acute risk period following booster doses and the post-acute period 28 days or more after the primary course, we observed increased incidence rates after the BNT1BNT2BNT3 and MOD1MOD2MOD3 schedules among males (Table 2). Among males, the largest IRR of 8.89 (95% CI, 2.26 to 35.03) was between MOD1MOD2MOD3 and MOD1MOD2. The IRR between BNT1BNT2BNT3 and BNT1BNT2 was 2.08 (95% CI, 1.31 to 3.33) (Table 2). These booster dose IRRs were largely unchanged compared to the booster dose comparisons with unvaccinated follow-up.

### Incidence rates after vaccination

Incidence rates observed in the 28-day acute risk periods following vaccination were larger among males than among females (Table 2). The rates were higher after a dose of MOD compared to after a dose of BNT, both in the case of a primary course and a booster dose. The incidence rates after a primary course were 6.87 cases (95% CI, 5.37 to 8.67) and 1.34 cases (95% CI, 1.08 to 1.63) in 28 days among 100,000 males after MOD and BNT, respectively. The incidence rates after a booster dose were 1.95 cases (95% CI, 0.53 to 4.99) and 0.86 cases (95% CI, 0.53 to 1.32) in 28 days among 100,000 males after MOD and BNT, respectively. The highest rates were observed in 16-to-24-year-old males after BNT1BNT2 (2.70 cases [95% CI, 2.06 to 3.47] in 28 days among 100,000 individuals) and after MOD1MOD2 (14.20 cases [95% CI, 9.51 to 20.39] in 28 days among 100,000 individuals). After BNT1BNT2BNT3, the highest rates were also observed in 16-to-24-year-old males (1.59 cases [95% CI, 0.79 to 2.85] in 28 days among 100,000 individuals). The use of MOD as a booster dose was restricted in younger males, and the incidence rate after MOD1MOD2MOD3 in 25-to-39-year-old males was 1.52 cases (95% CI, 0.31 to 4.45).

In 12-to-39-year-old females, the highest incidence rates were observed after MOD1MOD2 (0.37 cases [95% CI, 0.12 to 0.87] in 28 days among 100,000 individuals).

### Characteristics of myocarditis cases

Male cases were in general much more frequent independent of vaccination status (Table 3). In BNT vaccinated cases, the proportion of females ranged from 17.2% to 21.5%, and in MOD vaccinated cases, the proportion ranged from 0.0% to 25.4%. The cases occurring within the 28-day acute risk period occurred at a median of 5.6 and 7.2 days after the second and third BNT vaccinations, respectively, and 3.1 and 17.2 days after the second and third MOD vaccinations, respectively. Dosing intervals were similar across the different mRNA schedules, and the median length-of-stay in hospital ranged from 3.0-5.0 days (Table 3).

**Table 3:** Characteristics of myocarditis cases according to vaccination status at date of diagnosis among 12-to-39-year-olds in Denmark, Finland, Norway, and Sweden.

| Vaccination status<sup>1</sup> | Events | Females (%) | Number of days since last dose, median (IQR) | Dosing interval, median (IQR)<sup>2</sup> | Number of days spent in hospital, median (IQR) |
| --- | --- | --- | --- | --- | --- |
| BNT1BNT2 28-day acute risk period | 116 | 17.2% | 5.6 (3.7 to 10.9) | 46.7 (34.8 to 57.5) | 3.3 (2.4 to 4.3) |
| BNT1BNT2 post-acute risk period | 295 | 19.3% | 129.3 (79.9 to 203.1) | 53.3 (41.8 to 60.8) | 3.4 (2.3 to 5.0) |
| BNT1BNT2BNT3 28-day acute risk period | 25 | 16.0% | 7.2 (5.7 to 13.2) | 162.6 (150.8 to 196.6) | 3.9 (2.5 to 4.8) |
| BNT1BNT2BNT3 post-acute risk period | 65 | 21.5% | 110.8 (75.9 to 143.4) | 152.7 (144.1 to 183.9) | 3.4 (2.1 to 4.4) |
| MOD1MOD2 28-day acute risk period | 76 | 6.6% | 3.1 (2.9 to 6.7) | 50.2 (39.2 to 55.1) | 3.1 (2.4 to 4.0) |
| MOD1MOD2 post-acute risk period | 63 | 25.4% | 103.7 (69.7 to 153.5) | 49.9 (41.5 to 55.5) | 3.1 (2.0 to 4.0) |
| MOD1MOD2MOD3 28-day acute risk period | 4 | 0.0% | 17.2 (12.0 to 17.2) | 180.5 (171.5 to 186.5) | 5.0 (2.0 to 6.5) |
| MOD1MOD2MOD3 post-acute risk period | 3 | 0.0% | 124.7 (74.3 to 175.0) | 167.3 (164.7 to 170.0) | 3.0 (2.7 to 3.3) |
| Unvaccinated | 496 | 17.5% | NA (NA to NA) | NA (NA to NA) | 3.5 (2.3 to 5.1) |

<sup>1</sup>At time of myocarditis diagnosis.
<sup>2</sup>The number of days between first and second dose for two-dose schedules and the number of days between the second and third dose for three-dose schedules.

## DISCUSSION

We observed an acute increase in the risk of myocarditis in 12-to-39-year-old males following an mRNA booster vaccination, most prominently after MOD. However, this risk was attenuated compared to the risk after a primary course. In 12-to-39-year-old females, we also observed acute increased risk of myocarditis following mRNA vaccination. However, myocarditis was much rarer in females compared to males. Our results suggest that on average, 0.9 and 2.0 males aged 12–39 years will experience myocarditis hospitalization per 100,000 BNT and MOD booster vaccinated, respectively. Among 16-to-24-year-old males, 1.6 will experience hospitalization per 100,000 BNT booster vaccinated, and among 25-to-39-year-old males, 1.5 will experience hospitalization per 100,000 MOD booster vaccinated.

Few studies have evaluated the association between mRNA boosters and risk of myocarditis. One self-controlled case series study from the UK included 85 cases of myocarditis after a BNT booster and 8 after a MOD booster.^6^ Among males 13 to 39 years of age, a relative risk of 2.28 (95% CI, 0.77 to 6.80) was reported in the 28-day period after the BNT booster; no estimates were presented for the MOD booster in this group. The strength of this association compares well with our observations (IRRs of 2.21 [95% CI,1.37 to 3.57] and 2.08 [95% CI, 1.31 to 3.33], depending on our comparison group). However, our estimates of the association between a BNT booster and myocarditis (independent of comparison group) have higher statistical precision, and we were also able to provide estimates for the MOD booster. A French case-control study that included 31 myocarditis hospitalizations after the BNT booster, reported an odds ratio of 4.9 (95% CI, 3.1 to 7.7) among 12-to-29-year-olds.^7^ This is somewhat higher than the estimates observed in our study, but in a slightly younger age group and with a 7-day acute risk period compared to our 28-day acute risk period. Furthermore, no sex-stratified or MOD booster results were presented. In a comparison of post-vaccination incidence rates to historical rates of myocarditis conducted in a US study, a relative risk of 2.61 (95% CI, 1.13 to 5.29) was reported within a 21-day acute risk period after an mRNA booster dose based on 9 exposed cases.^5^ However, the study was conducted in a general population cohort with a median age of 47 years and a female proportion of 53.6%, and no results were presented separately for BNT or MOD. In another comparison of post-vaccination incidence rates and historical rates from British Columbia, Canada, a relative risk of 5.81 (95% CI, 3.83 to 8.46) was reported within a 21-day acute risk period after an mRNA booster based on 27 cases.^8^ Our study adds significantly to the current evidence from analytical studies on the association between mRNA boosters and myocarditis by providing age-, sex-, and vaccine type-specific results from nationwide cohorts in the Nordic setting with free universal healthcare and complete registration of vaccinations and myocarditis hospitalizations.

### Strengths and limitations

A key strength of our study is the use of two comparison follow-up periods: unvaccinated follow-up and post-acute risk period follow-up among primary course vaccinated. Previous analytical studies either compare with only unvaccinated or mix both unvaccinated and vaccinated in the comparison group. It could be argued that the comparison with primary course vaccinated follow-up is the most relevant comparison from a public health perspective in order to answer the following important research question: “What is the risk of myocarditis after the booster dose among individuals already vaccinated with a primary course?”. Furthermore, unvaccinated individuals may be a highly selected group in the Nordic populations, with high vaccine uptake (78%–87% vaccinated). However, unvaccinated follow-up time in our study mostly comprised follow-up time that occurred before the first dose among individuals who were later vaccinated, while individuals who have remained unvaccinated throughout the pandemic contributed only a small share of unvaccinated follow-up time.

Due to public awareness of the relationship between myocarditis and COVID-19 vaccination, milder cases may have been diagnosed more often in the period following vaccination compared to other time periods, which could lead us to overestimate the incidence rates and the IRRs of myocarditis after vaccination. This could be more pronounced after the booster, which temporally coincided with increased publicity and awareness. However, this potential bias was of limited concern, since our results were consistent also when the comparison was made to post-acute time following the primary course, which also occurred during periods of raised awareness. Another potential limitation is the lack of medical chart validation of myocarditis cases. However, the use of ICD-10 codes to ascertain myocarditis outcomes has previously been demonstrated to be sufficiently predictive for research purposes.^11^ In non-geriatric adults below 60 years of age, the positive predictive value of ICD-10 coded myocarditis was 85% in the Danish National Patient Registry. The misclassification would only affect our IRRs if it would differ between the risk period and the comparison period.

Our meta-analysis approach has the weakness that when pooling the adjusted IRR estimates, we did not utilize estimates from a country if there were no myocarditis events in either the risk or comparison periods. As the risk periods are shorter than the comparison periods, this exclusion is more likely to be due to no events in the risk period, which can lead to underrepresentation of the follow-up time during the risk period in the meta-analysis. This may, in turn, lead to an overestimation of the incidence rates of myocarditis during the acute risk periods and the IRRs.

## CONCLUSIONS

In conclusion, our results suggest that booster dose vaccination, just like the primary course vaccinations, is associated with increased myocarditis risk in the first 28 days following vaccination in male adolescents and young male adults. We estimate that 2.0 and 0.9 per 100,000 12-to-39-year-old males will experience myocarditis hospitalization within 28 days of MOD and BNT booster vaccination, respectively.

## Data Availability

The data comprises identifiable and sensitive individual-level health data which cannot be directly shared by us according to Danish law. However, it is possible, upon reasonable request, to obtain access to the underlying register data subject to Danish regulations for researcher access.

## Author Contributions

Dr Hviid and Mr Nieminen had full access to all of the data in the study and take responsibility for the integrity of the data and the accuracy of the data analysis.

*Concept and design*: Hviid, Nieminen, Gunnes, Karlstad, Hansen, Gulseth, Husby, Pihlström, Ljung, Hovi.

*Acquisition, analysis, or interpretation of data*: Hviid, Nieminen, Pihlström, Gunnes, Dahl, Karlstad, Gulseth, Sundström, Husby, Hansen, Ljung, Hovi.

*Drafting of the manuscript*: Hviid.

*Critical revision of the manuscript for important intellectual content*: Hviid, Nieminen, Pihlström, Gunnes, Dahl, Karlstad, Gulseth, Sundström, Husby, Hansen, Ljung, Hovi.

*Statistical analysis*: Nieminen, Pihlström, Gunnes, Hansen, Dahl, Sundström.

*Obtained funding*: Gulseth, Hviid.

*Administrative, technical, or material support*: Hviid, Karlstad, Gulseth, Ljung, Hovi.

